# Catastrophic Health Expenditure Among Gastrointestinal Cancer Patients Undergoing Surgery in Uganda: A Prospective Study

**DOI:** 10.64898/2026.03.18.26347930

**Authors:** Vivian Valin Akello, Clara Odhiambo Atieno, Lois Asiimwe, Gideon Kwikiriza Kurigamba, Mary Nakafeero, Kenneth Nkangi, Rachael Waako, Dickens Kamugisha, Urlich Vickos, Josephat Jombwe, Rosemary Byanyima

## Abstract

**Background:** The global burden of gastrointestinal (GI) cancers is projected to rise by 2050, with incidence and mortality in Africa nearly double global estimates. Surgery remains the cornerstone of treatment but imposes substantial financial burdens. In Uganda, where no national health insurance scheme exists, patients are especially vulnerable. We therefore investigated the magnitude of catastrophic health expenditure (CHE) among GI cancer patients undergoing surgery in public hospitals.

**Methods:** A prospective study was conducted over 10 months in the GI surgery wards of a tertiary hospital, with ethics approval. Adults with GI cancer scheduled for surgery were consecutively recruited. Sociodemographic, clinical, and household expenditure data were collected at baseline and discharge. Out-of-pocket (OOP) costs, annual household expenditure, non-food expenditure, and capacity to pay were derived. CHE was assessed using Wagstaff/van Doorslaer and Xu thresholds and determinants of CHE assessed.

**Results:** 164 participants were recruited, 54.3% were male and 75.0% aged above 50 years. The median out-of-pocket (OOP) expenditure for GI cancer surgery was USD 663, nearly twice the median annual household income. At the 10% threshold, the prevalence of CHE was 64%. Sources of financing for OOP varied by socioeconomic status (SES): households in the highest SES relied primarily on savings, whereas those in the lowest SES depended on asset sales and loans. School fees payment was disrupted, particularly among middle- and low-SES households. Factors independently associated with higher CHE included female sex, formal employment, curative intent of surgery, and low household SES.

**Conclusion:** Over half of patients experienced financial toxicity, often selling assets and compromising long-term security. The burden was greatest among poorer households, women, and those undergoing curative surgery. Findings highlight the urgent need for national health insurance in Uganda. Although recall bias may have influenced reporting, critical gaps in financial protection for cancer surgery patients are evident.

**Summary Box:** *What is already known about this topic:*

- Gastrointestinal cancers are rising in incidence across Africa yet treatment of GI cancers is costly due to the multimodal treatment approaches. GI cancer treatment and surgery lead to catastrophic health expenditures even in high income countries.

*What this study adds:*

- This study evaluates surgery as a key management modality for gastrointestinal cancers and quantifies the catastrophic health expenditure associated with it, found to be **64%**.
- It identifies risk factors for CHE in this context, emphasizing the vulnerability of households undergoing surgical cancer care.
- It highlights differences in sources of health care financing across socioeconomic strata, revealing inequities in how households mobilize funds.
- It highlights basic needs that are negatively affected by the shortage of resources such as education, and reveals a high likelihood of future financial hardship due to the impact of crowding out effect on income generating activities.

*How might this impact on clinical practice?:*

- Surgery remains one of the most cost-effective and potentially curative modalities for gastrointestinal cancers, these findings stress the urgent need for financial risk protection strategies in Uganda.
- National cancer plans should prioritise procurement of specialised surgical equipment and safe guard vulnerable individuals especially females and financially deprived who stand to benefit from curative surgery.

## Background

Two thirds of global deaths are caused by non-communicable diseases(NCDs), about three quarters of these deaths occur in low and middle income countries(LMICs), accounting for 86% of premature mortality in these countries(1, 2). Cancers are the second most prevalent NCD worldwide, demographic shifts including population growth, longer life expectancy, urbanization and adoption of new lifestyles are some of the factors that are driving a rise of incidence especially in Africa(3). By 2030 it is projected that there will be about 70% increase in cancer cases and mortality associated with cancers globally, however in Africa these figures are likely to exceed the global rates by 30%(4).

Gastrointestinal Cancers (GI) cancers represent a major share of this burden, representing one in four cases and one in three cancer related deaths (5). GI cancer incidence is projected to rise by 83% and mortality by 93% by 2050, with Africa experiencing nearly double the global rate(6). And yet, more than half of GI cancers are attributable to modifiable risk factors, which include smoking, alcohol use, metabolic syndromes, infections, and dietary patterns-which vary across cultures and geographies highlighting the potential for preventive intervention (7). The financial impact of cancer care is profound, affecting health systems, patients and families even in settings of financial protection systems (8).

The highest 5-year survival rates, up to 80% in GI cancers is noted in patients who are treated with surgery alone or in combination with chemotherapy or radiotherapy(9). Therefore, surgery offers patients with GI cancer the highest chance of long-term survival. GI cancer surgery is complex and resource intensive, often requiring prolonged hospitalisation and carrying high risks of postoperative complications that escalate costs. In Africa, out-of-pocket (OOP) payments account for approximately 25% of healthcare expenditures, rising to over 40% in LMICs often leading to catastrophic health expenditure (CHE) and impoverishing health expenditure (IHE) (10, 11).

In SSA, CHE is estimated at 16.5% for a 10% threshold, appears to have a rising trend over the years, with higher estimates noted among households affected by cancer and surgical populations(10, 12). These financial shocks force households to borrow, sell assets, or forgo treatment, with long-term consequences including indebtedness and crowding-out effects with resource displacement from key areas like education. (3, 12, 13). These dynamics undermine human capital development and perpetuate generations of poverty.

Financial protection is a central goal of Sustainable Development Goal(SDG) 3.8 which entails achievement of universal health coverage ensuring access to quality health services without financial hardship or threats to living standards(11). Social protection mechanisms—including tax-based financing, social health insurance, and community-based schemes—are critical to reducing financial barriers(14). Yet, coverage remains low in LMICs, with health insurance penetration below 8% and weak social support systems(3, 14). Surgical care remains grossly underfunded, with no African country allocating a dedicated budget line for surgery—further compounding the challenges faced by GI cancer patients across the continent (15).

In Uganda, the absence of a national health insurance scheme exacerbates financial vulnerability in healthcare. Private insurance remains limited to formal sector employees and wealthier individuals, with national coverage at only 1.1% (16). OOP payments are widespread, affecting one-third of surgical patients even in public institutions, with 29% experiencing CHE and 31% incurring IHE (17). These figures are likely higher among cancer patients requiring specialised GI surgery. Stock-outs of essential medicines, the need for specialised equipment and drugs, radiological investigations, and postoperative critical care further drive substantial OOP costs for GI cancer procedures. Such payments disproportionately burden the poor, undermining Uganda’s national priority of poverty eradication.

Despite the rise in GI cancers and the already known substantial OOP experienced by surgical population and the lack of a functional social and financial protection mechanism in public health facilities, evidence on the extent of financial distress among GI cancer patients remains limited. We therefore set out to investigate the magnitude of CHE among gastrointestinal ancer patients undergoing surgery in a free-to-access public hospital and to evaluate the factors associated with CHE in this population.

## METHODS

### Study Design and Setting

We conducted a prospective observational study over 10 months with two data collection points at baseline (preoperative) and discharge. Recruitment occurred between February and October 2024 in the gastrointestinal surgery wards of Mulago National Referral Hospital (Upper GI and Hepato-pancreatobiliary, Colorectal, and Cardiothoracic wards). Eligible participants were adults (≥18 years) admitted for planned GI cancer surgery and consent to inclusion in the study. Patients who had recurrent cancers surgeries were excluded from the study. Consecutive sampling was used to enrol respondents. Written informed consent was obtained from all participants, and ethical approval was granted by the Mulago Hospital Research and Ethics Committee. Patients and the public were not involved in the design, conduct, reporting or dissemination of this study.

### Data Collection

Interviewer-administered questionnaires captured patient biodata (age, sex, education, employment, residence), diagnosis, and surgical procedure (curative or palliative). Household expenditure data were collected at baseline and discharge, including monthly spending on food, healthcare, transport, donations, clothing, home improvements, education, and other items. Hospitalisation-related expenditures comprised transport, attendant costs, pre-operative medication, surgical supplies, investigations, intensive care unit (ICU) admission, and informal payments. Sources of funds for hospitalisation were also documented. Patients who died after surgical intervention were included, as the bulk of surgical costs are incurred on the day of surgery and no loss to follow up occurred since patients were followed up to discharge.

### Data Management

Data were electronically captured using Kobo Collect with programmed logical skips and range checks for quality assurance and this minimised entry errors. All recruited participants had complete datasets; no missing data were encountered. Data were exported to Microsoft Excel for monitoring, then cleaned and analysed in Stata version 15. Baseline and discharge records were merged using unique patient identifiers. Annual household expenditure was estimated by multiplying average monthly expenditure by 12. Non-food expenditure was calculated by excluding food costs. Hospitalisation-related out-of-pocket (OOP) expenditure was derived from transport, attendant costs, medication, surgical supplies, investigations, and other medical expenses. Household capacity to pay was defined as monthly expenditure excluding food, transport, and clothing.

### Measurement of Catastrophic Health Expenditure (CHE)

CHE incidence was assessed using two approaches:

1. Wagstaff and van Doorslaer method: OOP health payments exceeding 10% of total expenditure or 40% of non-food expenditure.
2. Xu et al. method: OOP payments exceeding 40% of household capacity to pay.

CHE headcount was defined as the proportion of households incurring catastrophic expenditure. Intensity was measured using the overshoot, representing the average degree by which hospitalisation expenditures exceeded the threshold. The mean positive overshoot (MPO) was calculated as overshoot divided by headcount.

Data were collected on sources of funds used to meet out-of-pocket payments, as well as on household projects whose financing was displaced.

### Statistical Analysis

Descriptive statistics included frequencies and percentages for categorical variables, and means, medians, standard deviations, or interquartile ranges (IQRs) for continuous variables.

Logistic regression was used to explore pairwise associations between CHE at the 10% threshold and the independent variables. Multicollinearity was ruled out using the variance inflation factors (VIF) approach and the highest VIF value was 2.1.

Backward elimination was employed to identify predictors of CHE, retaining variables significant at the 5% level in the final parsimonious multivariable model. The Hosemer-Lemeshow goodness of fit-test had a p-value of 0.8521 implying that the final model fitted the data well.

### Sensitivity Analysis

CHE thresholds were varied to test robustness. Analyses were stratified by cancer type and income quintiles to explore heterogeneity.

## RESULTS

The sociodemographic and economic characteristics of respondents is shown in Table 1 above. A total of 164 respondents were recruited in the study. Just over half were male (54.3%), with a mean age of 54.3 years; notably, 75% of the participants were aged 50 years and above. The majority were employed in the informal sector (77.4%) and resided in rural areas (56.7%). The median annual household expenditure was 1,242,500 Uganda shillings (≈345 USD), while the median out-of-pocket (OOP) health expenditure reached 2,385,000 shillings (≈663 USD). In contrast, the median household capacity to pay was only 620,000 shillings (≈172 USD). These figures underscore the disproportionate burden of OOP costs relative to household resources, highlighting the socioeconomic vulnerability faced by patients with gastrointestinal cancers.

**Table 1:** Sociodemographic and Economic Characteristics of Gastrointestinal Cancer Patients at Mulago National Referral Hospital (n=164)

| **Characteristic** | **n (%) / Median [IQR]** | **Range** |
| --- | --- | --- |
| **Sex** | Male: 89 (54.3), Female: 75 (45.7) | – |
| **Age (years)** | Mean (SD): 57.9 (13.6); Median [IQR]: 59.5 [49.5–68.5] | 24–94 |
| **<50 years** | 41 (25.0) | – |
| **≥50 years** | 123 (75.0) | – |
|  | 18 (10.9) | – |
| **Education** | Primary: 96 (58.5), Secondary: 40 (24.4), Tertiary: 28 (17.1) | – |
| **Employment sector** | Informal: 127 (77.4), Formal: 37 (22.6) | – |
| **Residence** | Urban: 71 (43.3), Rural: 93 (56.7) | – |
| **Annual household expenditure** | 1,242,500 UGX [600,000–2,880,000]<br>(≈330 USD [160–760]) | 60,000–14,500,000 UGX<br>(≈16–3,850 USD) |
| **OOP health expenditure** | 2,385,000 UGX [1,219,350–4,795,000]<br>(≈630 USD [320–1,270]) | 841,000–32,500,000 UGX<br>(≈220–8,630 USD) |
| **Capacity to pay** | 620,000 UGX [170,000–1,760,000]<br>(≈165 USD [45–470]) | 100,000–10,800,000 UGX<br>(≈27–2,870 USD) |

*Conversion based on average exchange rate: 1 USD ≈ 3,750 UGX (2024). OOP is out-of-pocket.*

Overall, 45.2% of respondents underwent surgery with curative intent, while the remainder received palliative procedures. Among the informally employed, only 34.6% accessed potentially curative surgery, compared with 81.1% of those formally employed.

At the 10% threshold of annual household expenditure, 64% of households experienced CHE. This proportion dropped at higher thresholds, as shown in Table 2. On average, households exceeded the 10% threshold by 25.9 percentage points, and among those affected, the mean overshoot was 40.4 percentage points.

**Table 2.** Intensity of Catastrophic Health Expenditure among GI Cancer Surgery Patients at Mulago National Referral Hospital.

| **Expenditure Definition** | **Measure13** | **10% Threshold** | **15% Threshold** | **25% Threshold** | **40% Threshold** |
| --- | --- | --- | --- | --- | --- |
| **Total Expenditure** | Headcount (%) | 64.0<br>(105/164) | 53.7<br>(88/164) | 35.9<br>(59/164) | – |
|  | Overshoot (%) | 25.9 | 22.9 | 18.7 | – |
|  | Mean positive overshoot (%) | 40.4 | 42.8 | 52.1 | – |
| **Non-food Expenditure** | Headcount (%) | – | 61.6<br>(101/164) | 45.1<br>(74/164) | 31.7<br>(52/164) |
|  | Overshoot (%) | – | 76.6 | 71.4 | 49.2 |
|  | Mean positive overshoot (%) | – | 124.3 | 158.2 | 155.2 |
| **Capacity to** | Headcount (%) | – | – | 60.4 | 48.2 |
| **Pay** |  |  |  | (99/164) | (79/164) |

When non-food expenditure was used as the denominator, both incidence and intensity were greater. At the 40% threshold, the CHE headcount was 31.7%, with overshoot values of 49.2 percentage points. The mean positive overshoot exceeded 100 percentage points across all thresholds, reflecting substantial financial distress among affected households.

Using capacity to pay as the expenditure definition, the incidence of CHE remained high at 60.4%. Overall, increasing the threshold reduced the proportion of households crossing it, but among those that did, the financial burden was considerably heavier.

When stratified by socioeconomic status, distinct patterns emerge in how households financed surgery as shown in Figure 1. Overall, all households relied on funding from friends and family (68.3%) with less dependence on savings, loans or sale of assets. High-SES households predominantly relied on savings (35.2%) and support from family and friends, with minimal dependence on loans and no reported asset sales. Middle-SES households largely depended on contributions from family and friends (80%), supplemented by loans, savings, and occasional asset sales. Low-SES households showed the most precarious strategies, frequently resorting to asset sales (23.6%) and loans (14.6%), with limited reliance on savings (3.6%).

**Fig 1:**
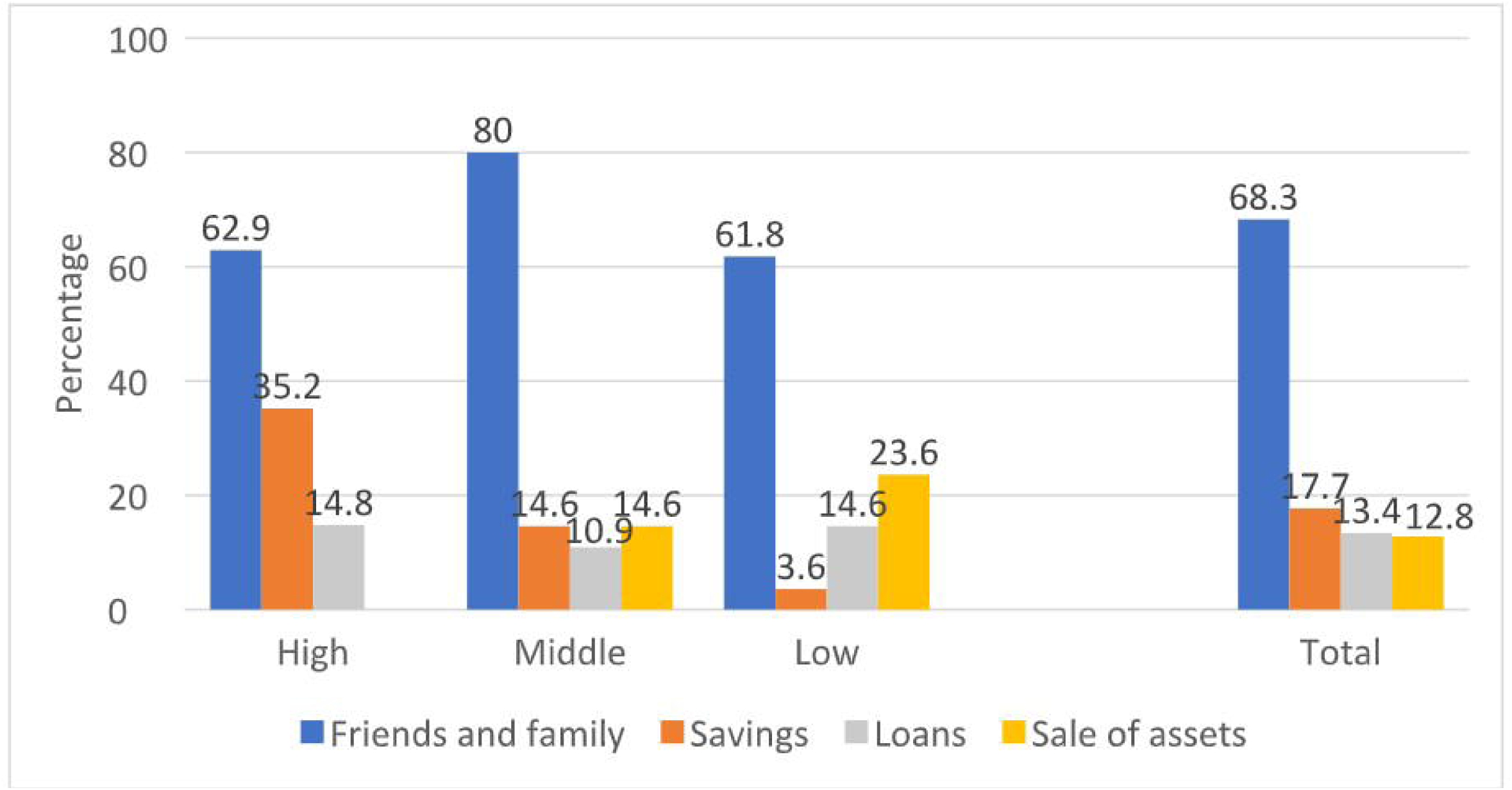
Sources of Out-of-Pocket Health Expenditure by Socio-economic Status

These findings highlight the unequal coping mechanisms across socioeconomic strata, underscoring the vulnerability of lower-income households that must liquidate assets or incur debt to meet the costs of cancer surgery.

Catastrophic health expenditure produced notable crowding-out effects on household priorities as shown in Figure 2. Overall, family businesses were most affected (33.5%) followed by school fees (23.8%). Family business investments were disproportionately curtailed among high-SES households (44.4%).). Family businesses and school fees were affected in similar magnitudes in the middle-SES.

**Fig 2:**
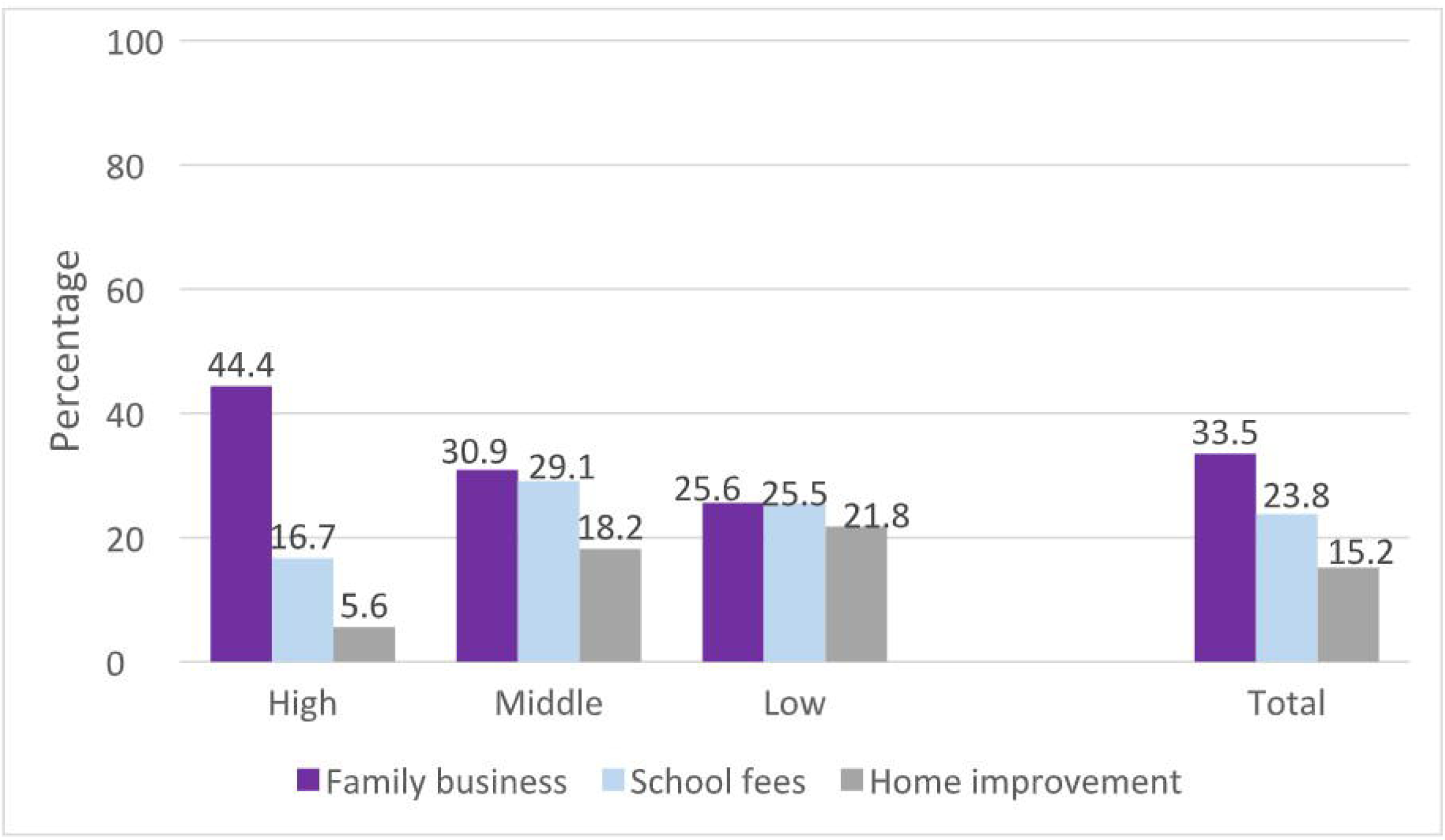
Crowding-Out Effects of Catastrophic Health Expenditure (CHE) on Household Projects by Socioeconomic Status

At least one in every 5 households in the low-SES reported crowding out effect in all the three areas, with home improvement projects observed to be more affected (21.8%) when compared to other socio-economic statuses.

These patterns illustrate how CHE disrupts essential and developmental household projects across socioeconomic strata, with lower-income households particularly vulnerable to trading off long-term investments in education and housing.

Multivariable regression identified several sociodemographic and clinical predictors of CHE as shown in Table 3. Female respondents were nearly five times more likely to incur CHE compared with males, (aOR=4.96, 95% CI:2.19,11.27, p<0.001). Individuals engaged in informal employment were significantly less likely to experience CHE than those in formal employment, (aOR=0.29, 95% CI: 0.09,0.83, p=0.021). Household wealth status showed a strong, graded association: compared with respondents from high-wealth households, those in middle socioeconomic status were six times more likely, CHE (aOR=5.89, 95% CI: 2.16,16.09, p=0.001), and those in low socioeconomic status fourteen times more likely, to experience CHE (aOR=13.86, 95% CI: 4.35,11.08, p<0.001). Finally, patients undergoing curative surgery were almost five four times more likely to incur CHE compared with those receiving palliative surgery, (aOR=3.95, 95% CI: 1.57,9.88, p=0.003). These findings expose how the effects of CHE affect the most vulnerable in society, women and financially deprived.

**Table 3:** expenditure among gastrointestinal surgery patients at Mulago National Referral Hospital.

| **Variable** | **Proportion of OOP exceeding 10% (%)** | **Crude OR (95% CI)** | **Adjusted OR (95% CI)** | **p-value** |
| --- | --- | --- | --- | --- |
| **Overall (n=164)** | 64.0 | – | – | – |
| **Sex** |  |  |  |  |
| **Male (n=89, ref)** | 53.9 | 1.00 | 1.00 | – |
| **Female (n=75)** | 76.0 | 2.71 (1.38–5.31) | 4.96 (2.19–11.27) | <0.001 |
| **Age** |  |  |  |  |
| **≥50 years (n=123, ref)** | 66.7 | 1.00 | – | – |
| **<50 years (n=41)** | 56.1 | 0.64 (0.31–1.32) | – | – |
| **Education level** |  |  |  |  |
| **Tertiary (n=28, ref)** | 75.0 | 1.00 | – | – |
| **Secondary (n=40)** | 52.5 | 0.37 (0.13–1.06) | – | – |
| **Primary (n=96)** | 65.6 | 0.64 (0.25–1.65) | – | – |
| **Sector employed** |  |  |  |  |
| **Formal (n=37, ref)** | 73.0 | 1.00 | 1.00 | – |
| **Informal (n=127)** | 61.4 | 0.59 (0.26–1.32) | 0.29 (0.09–0.83) | 0.021 |
| **Residence** |  |  |  |  |
| **Urban (n=71, ref)** | 56.3 | 1.00 | – | – |
| **Rural (n=93)** | 69.9 | 1.79 (0.94–3.43) | – | – |
| **Wealth status** |  |  |  |  |
| **High (n=54, ref)** | 50.0 | 1.00 | 1.00 | – |
| **Middle (n=55)** | 69.1 | 2.24 (1.02–4.89) | 5.89 (2.16–16.09) | 0.001 |
| **Low (n=55)** | 72.7 | 2.67 (1.20–5.92) | 13.86 (4.35–44.08) | <0.001 |
| **Length of stay (days)** | Mean (SD)<br>20.1 (18.7) | 0.99 (0.98–1.01) | – | – |
| **Primary diagnosis** |  |  |  |  |
| **Esophageal cancer (yes vs. no)** | 54.7 vs.<br>68.5 | 0.56 (0.28–1.09) | – | – |
| **Gastroduodenal cancer (yes vs. no)** | 61.1 vs.<br>64.8 | 0.85 (0.39–1.83) | – | – |
| **Hepatopancreatobiliary cancer (yes vs. no)** | 73.7 vs.<br>62.8 | 1.66 (0.57–4.87) | – | – |
| **Colorectal cancer (yes vs. no)** | 71.4 vs.<br>60.2 | 1.65 (0.83–3.32) | – | – |
| **Surgical intent** |  |  |  |  |
| **Palliative (n=90, ref)** | 56.7 | 1.00 | 1.00 | – |
| **Curative (n=74)** | 73.0 | 2.07 (1.06–3.99) | 3.95 (1.57–9.88) | 0.003 |

*Ref- Reference value used in analysis.*

Multivariable regression identified several sociodemographic and clinical predictors of CHE as shown in Table 3. Female respondents were nearly five times more likely to incur CHE compared with males. Individuals engaged in informal employment were significantly less likely to experience CHE than those in formal employment. Household wealth status showed a strong, graded association: compared with respondents from high-wealth households, those in middle socioeconomic status were six times more likely, and those in low socioeconomic status fourteen times more likely, to experience CHE. Finally, patients undergoing curative surgery were almost five times more likely to incur CHE compared with those receiving palliative surgery. This findings expose how the effects of CHE affect the most vulnerable in society, women and financially deprived.

## DISCUSSIONS

The rate of catastrophic health expenditure in patients undergoing GI cancer surgery was 64% for 10 % threshold. Our estimate is slightly lower than that reported in a recent LMIC-focused review, which found a pooled prevalence of catastrophic health expenditure (CHE) of 71.96% for gastrointestinal cancers at the 10% threshold—substantially higher than the pooled prevalence of 56.89% for all cancers at the same threshold, though this review was not restricted to a single treatment modality (18). Another review reported a CHE prevalence of 59% among GI cancer patients, attributed to multiple costly treatment modalities; although lower than our estimate, this analysis also encompassed non-surgical treatments (19). CHE is consistently greater in lower-income countries, largely due to weak social and financial protection mechanisms (19). Treatment costs for GI cancers are higher than for other cancers across both high- and low-income settings, and surgery especially in a low resource setting adds to this financial burden as shown by this study. Yet, compared with chemotherapy, hormonal therapy, radiotherapy, and palliative care, surgery remains the most cost-effective (20). This study provides a unique lens on gastrointestinal cancer surgeries as a single specific treatment modality and their economic impact on patients in a low-income country. Ensuring financial protection for surgical patients is therefore critical—not only to safeguard individuals but also to strengthen the resilience of health systems. Findings from our study reinforce the established evidence that surgery increases the risk of CHE in cancer patients (21).

### Coping Mechanisms of OOP

In our study, median out-of-pocket (OOP) expenditure for GI cancer surgery exceeded annual household expenditure, resulting in potentially severe financial hardship and long-term economic consequences. Cancer care imposes financial toxicity, encompassing OOP payments and the psychological distress, and coping behaviours associated (20). Patients without social protection commonly rely on crowding funding, relying on family, friends, and workplaces for financial support and at the individual level, coping strategies include depleting savings, selling assets, incurring debt, or foregoing treatment(20, 22, 23). This finding was observed in this study, with community and family support as the most frequent coping mechanisms across all socioeconomic strata. Coping mechanisms varied by income level. High-income earners primarily relied on depleting savings, which helped protect them from selling property. In contrast, low- and middle-income groups depended more heavily on loans and asset sales which included land and livestock. Among the lowest-income earners, reliance on savings was minimal—likely because such reserves were unavailable—leading to greater asset liquidation.

Families also displaced funds (crowding out) from essential areas such as income generation, home improvement, and school fees. Education was most affected, particularly in low- and middle-income households, despite its importance for escaping poverty. Similar findings have been reported elsewhere(13). This displacement undermines future income generation and long-term prosperity, with school dropouts more likely among low- and middle-income families who often fail to recover from the financial catastrophe.

### Factors associated with Catastrophic Health Expenditure

The risk of CHE in our study was associated with socioeconomic status, gender, employment, and curative intent of surgery. Wealth status is a well-established determinant of CHE, with higher socioeconomic status offering protection(18, 19, 24). Patients with lower wealth levels lack adequate savings and social support mechanisms, leaving them more exposed to financial toxicity. They often resorted to distressing financing like loans, sale of assets or diversion of funds from essential needs to cover out-of-pocket costs, further destabilizing their financial future. Measures to protect the financially disadvantaged from further impoverishment especially while accessing health care are vital to eradicate poverty.

Female patients were disproportionately affected by catastrophic health expenditure (CHE). In Uganda, fewer women hold formal employment, and those who do are often concentrated in lower-cadre positions(25). Moreover, these women have less access to education, stable work opportunities, and financial resources (26). They also shoulder the bulk of unpaid domestic labor, which entrenches poverty and reinforces dependence on others for health-related financial decisions (26). This similar gendered vulnerabilities have been documented in other LMICs that has shown that women are financially dependent on men causing lack of autonomy in medical decision making (18).These intersecting structural disadvantages compound women’s exposure to CHE and highlight the urgent need for gender-sensitive health financing policies.

While most studies suggest that formal employment is protective against CHE, our findings diverge(18). In Uganda, the informal sector is not subject to income tax, standard labour legislation, or employee benefits(25). Only about one third of informally employed patients accessed surgery with a curative intent. 90% of Ugandans are informally employed, these findings show that the bulk of the population are left out of potentially curative surgery possibly due to delays in seeking care and diagnosis. Palliative procedures are less resource-intensive, associated with shorter hospital stays, and require fewer investigations and consumables. As a result, they generate lower out-of-pocket expenditure and reduce the risk of catastrophic health expenditure (CHE) compared with curative surgical management—explaining the deviation observed.

Curative gastrointestinal oncological surgery was strongly associated with catastrophic health expenditure (CHE). These procedures typically require extensive preoperative investigations, specialized consumables, and often admission to intensive or high-dependency units due to there complexity. Even within public hospitals, cost-sharing policies and frequent equipment stock-outs substantially increase patient expenses. Longer hospital stays and higher complication rates further compound the financial burden. Evidence from a multicountry study in sub-Saharan Africa demonstrated that ICU admission, prolonged hospitalisation and complex surgeries were significant drivers of CHE among families of children requiring surgery (22).

Paradoxically, patients undergoing curative surgery—those with the greatest chance of long-term survival—remain burdened by ongoing costs of chemotherapy, surveillance imaging, and repeated investigations. This perpetuates financial toxicity, undermining recovery and long-term wellbeing. Although previous studies reported high CHE among patients receiving palliative care in LMICs, our study specifically assessed palliative surgery (e.g, gastrostomy, ostomy, bypass procedures, or biopsy), which typically requires shorter admissions and fewer resources(27).

### Recommendations

National surgical plans must prioritize procurement of specialized equipment and adoption of new technologies essential for complex cancer surgeries (28).At the hospital level, scarce equipment should be allocated to patients most likely to benefit from curative procedures, while waiver policies should target low-income patients, particularly women. At the community level, advocacy for social and financial support is critical. Above all, a national health insurance scheme explicitly incorporating cancer care is urgently needed to alleviate financial burden and promote equitable access to life-saving surgical interventions. Uganda still relies heavily on out of pocket payments up to 29% to fund the health services in the country and this will alleviate the CHE associated(29).

The causes of late presentation, which result in a higher proportion of palliative surgical procedures among Ugandan patients, warrant further investigation. Addressing these factors is essential to improve timely access to specialised GI cancer surgeries.

## LIMITATIONS

This study has some limitations. First, monthly expenses were documented through patient recall, which is prone to bias, particularly under stressful circumstances such as illness. Second, we did not follow patients after discharge to capture costs related to postoperative complications potentially leading to an underestimation of catastrophic health expenditure (CHE). Relatively wide confidence intervals for the odds ratios were observed due to the small sample size. Finally, the study was conducted in surgical wards of a public referral hospital in a low-income country and focused on patients scheduled for surgery, which may limit the generalisability of findings to the wider population.

## CONCLUSION

Catastrophic health expenditure (CHE) among patients undergoing gastrointestinal cancer surgery is alarmingly high at 64%. The burden falls disproportionately on financially disadvantaged groups, vulnerable women, and those undergoing curative procedures. To cope, patients rely on social networks or resort to exhausting savings, incurring debt, and selling assets—strategies that erode long-term financial security. Less financially stable households are forced into more damaging coping mechanisms, such as asset liquidation and debt accumulation, further entrenching vulnerability.

Waiver policies that prioritize disadvantaged patients and a national health insurance scheme explicitly covering cancer care are urgently needed. Such measures would provide essential social protection, reduce financial distress, and promote equitable access to life-saving surgical interventions. Further studies are needed to investigate the reasons behind the higher rate of palliative surgeries, particularly among informally employed patients.

## Data Availability

All data produced in the present study are available upon reasonable request to the authors.

## Funding

This study was supported by the Mulago National Referral Hospital.

## Notes

### Competing Interest Statement

The authors have declared no competing interest.

### Author Declarations

Ethics committee of Mulago National Referral Hospital gave approval for this work.

## References

1. Anderson S. WHO Unveils ‘Invisible Numbers’ Of The NCD Crisis As Leaders Meet At United Nations - Health Policy Watch 2022. Available from: https://healthpolicy-watch.news/who-unveils-invisible-numbers-of-the-ncd-crisis-as-leaders-meet-at-un/.

2. Bigna JJ, Noubiap JJ. The rising burden of non-communicable diseases in sub-Saharan Africa. The Lancet Global Health. 2019;7(10):e1295–e6.

3. Odunyemi A, Islam MT, Alam K. The financial burden of noncommunicable diseases from out-of-pocket expenditure in sub-Saharan Africa: a scoping review. Health Promot Int. 2024;39(5).

4. Hamdi Y, Abdeljaoued-Tej I, Zatchi AA, Abdelhak S, Boubaker S, Brown JS, et al. Cancer in Africa: The Untold Story. Front Oncol. 2021;11:650117.

5. Arnold M, Abnet CC, Neale RE, Vignat J, Giovannucci EL, McGlynn KA, et al. Global Burden of 5 Major Types of Gastrointestinal Cancer. Gastroenterology. 2020;159(1):335–49 e15.

6. Danpanichkul P, Pang Y, Tothanarungroj P, Dejvajara D, Kim D, Saokhieo P, et al. Gastrointestinal cancer statistics in 2022 and projection to 2050: GLOBOCAN estimates across 185 countries Cancer. 2026;132(1).

7. Danpanichkul P, Suparan K, Tothanarungroj P, Dejvajara D, Rakwong K, Pang Y, et al. Epidemiology of gastrointestinal cancers: a systematic analysis from the Global Burden of Disease Study 2021. Gut. 2024;74(1):26–34.

8. Alzehr A, Hulme C, Spencer A, Morgan-Trimmer S. The economic impact of cancer diagnosis to individuals and their families: A Systematic review. Supportive Care in cancer. 2022;30:6385–404.

9. Arjun S. Global burden of five major types of gastrointestinal cancer. Prz Gastroenterol. 2024;19(3):236–54.

10. Mengistie CT, Girma SM, Mengistie BT, Yigzaw HD, Tadesse SZ, Estifanos H, et al. Financing surgery in LMICs: building sustainable and equitable systems for universal surgical access - a narrative review. BMC Surg. 2025;25(1):516.

11. Africa WHOROf. Trends in Financial Hardship Due to Out-of-Pocket Health Spending in the WHO African Region. Brazzaville, Republic of Congo; 2023.

12. Eze P, Lawani LO, Aguc UJ, Acharya Y. Catastrophic health expenditure in sub-Saharan Africa: systematic review and meta-analysis Bull World Health Organ. 2022;100:337–51J.

13. Houeninvo HG, Quenum VCC, Senou MM. Out- Of- Pocket health expenditure and household consumption patterns in Benin: Is there a crowding out effect? Health Econ Rev. 2023;13(1):19.

14. Eze P, Ilechukwu S, Lawani LO. Impact of community-based health insurance in low-and middle-income countries: A systematic review and meta-analysis. PLoS One. 2023;18(6):e0287600.

15. Ifeanyichi M, Aune E, Shrime M, Gajewski J, Pittalis C, Kachimba J, et al. Financing of surgery and anaesthesia in sub-Saharan Africa: a scoping review. BMJ Open. 2021;11(10):e051617.

16. (UBOS) UBoS. National Population and Housing Census 2024: Final Report, Volume I (Main Report). Kampala, Uganda: Uganda Bureau of Statistics; 2024.

17. Nwanna-Nzewunwa O, Oke R, Agwang E, Ajiko MM, Yoon C, Carvalho M, et al. The societal cost and economic impact of surgical care on patients’ households in rural Uganda; a mixed method study. BMC Health Serv Res. 2021;21(1):568.

18. Kaso AW, Regesu AH, Haftu HK, Agero G, Jima GH, Kaso T, et al. Magnitude of catastrophic health expenditure and its determinants among cancer patients in low and middle-income countries: a systematic review and meta-analysis. BMC Health Serv Res. 2025;25(1):1533.

19. Kitaw TA, Tilahun BD, Zemariam AB, Getie A, Bizuayehu MA, Haile RN. The financial toxicity of cancer: unveiling global burden and risk factors - a systematic review and meta-analysis. BMJ Glob Health. 2025;10(2).

20. Donkor A, Atuwo-Ampoh VD, Yakanu F, Torgbenu E, Ameyaw EK, Kitson-Mills D, et al. Financial toxicity of cancer care in low- and middle-income countries: a systematic review and meta-analysis. Support Care Cancer. 2022;30(9):7159–90.

21. Kasahun GG, Gebretekle GB, Hailemichael Y, Woldemariam AA, Fenta TG. Catastrophic healthcare expenditure and coping strategies among patients attending cancer treatment services in Addis Ababa, Ethiopia. BMC Public Health. 2020;20(1):984.

22. Yap A, Olatunji BT, Negash S, Mweru D, Kisembo S, Masumbuko F, et al. Out-of-pocket costs and catastrophic healthcare expenditure for families of children requiring surgery in sub-Saharan Africa. Surgery. 2023;174(3):567–73.

23. Udayakumar S, Solomon E, Isaranuwatchai W, Rodin DL, Ko YJ, Chan KKW, et al. Cancer treatment-related financial toxicity experienced by patients in low- and middle-income countries: a scoping review. Support Care Cancer. 2022;30(8):6463–71.

24. Edeh HC. Exploring dynamics in catastrophic health care expenditure in Nigeria. Health Econ Rev. 2022;12(1):22.

25. Guloba M. Gender Disparities in Social Protection in Uganda. 2019.

26. Programme UND. NDC Support Programme Uganda gender analysis. New York: UNDP; 2021.

27. Reid E, Ghoshal A, Khalil A, Jiang J, Normand C, Brackett A, et al. Out-of-pocket costs near end of life in low- and middle-income countries: A systematic review. PLOS Glob Public Health. 2022;2(1):e0000005.

28. Are C, Murthy SS, Sullivan R, Schissel M, Chowdhury S, Alatise O, et al. Global Cancer Surgery: pragmatic solutions to improve cancer surgery outcomes worldwide. Lancet Oncol. 2023;24(12):e472–e518.

29. Ministry of Health U. Annual Health Sector Performance Report 2023/24. Kampala, Uganda: Ministry of Health, Uganda; 2024.

